# Attitudes of chronic pain patients on long-term opioid therapy toward opioid tapering

**DOI:** 10.1101/2023.12.19.23300217

**Authors:** R. Jabakhanji, F. Tokunaga, G. Rached, A.D. Vigotsky, J. Griffith, T.J. Schnitzer, A. V. Apkarian

## Abstract

The chronic pain and opioid addiction epidemics interact with each other, potentially exacerbating each respective condition. Despite having modest efficacy, millions of chronic pain patients in the USA continue to use opioids as their primary source of pain management. The Centers for Disease Control recommends opioid tapering to diminish the risk of opioid dependence in chronic pain patients. However, tapering, even with physician oversight, can introduce additional harm. Thus, many pain clinicians remain ambivalent about undertaking opioid tapering. Here, we surveyed attitudes on the topic from the viewpoint of chronic pain patients who have been consuming opioids over long durations. We queried 127 chronic pain patients (pain duration = 13.5 ± 9.6 years) on long-term opioids (10.3 ± 8.2 years), primarily consuming hydrocodone or oxycodone. Sixty-six percent of participants were “very” or “extremely” interested in participating in an opioid tapering study. Patients emphasized the importance of controlling their pain during opioid tapering, and over 50% were also worried about craving symptoms. Both the desire for tapering and the worry of pain control were more pronounced in participants with a higher magnitude of ongoing back pain. The study demonstrates that most chronic pain patients using opioids are interested in decreasing opioid consumption. Yet, they worry about losing control of their chronic pain. These results imply patient-physician strategies that may aid the engagement of both parties in opioid tapering.

## Introduction

Opioids remain the most common treatment for chronic pain. Even though opioid prescription dispensation in the U.S. has steadily declined in the last 20 years [1-3]—in 2019, 22.1% of U.S. adults with chronic pain reported using a prescription opioid in the previous three months [4, 5]. Pain and analgesic opioid prescriptions [6, 7] constitute a recognized entry point toward opioid misuse, dependence, and overdose [8]. There are conflicting views and lines of evidence concerning opioids’ efficacy for the treatment of chronic pain [9-17]. Notwithstanding these divergent viewpoints, if and how to taper patients’ opioids remains a relatively unexplored topic.

To combat the opioid epidemic, in 2016, the CDC issued guidelines for decreasing opioid prescriptions nationwide [18]. In 2022, the CDC issued revised and more tempered guidelines, emphasizing patient-physician relationships regarding tapering decisions but still warning of opioids’ adverse effects [19]. These guidelines reduced rates of opioid dispensation [2, 3, 20, 21] but, perhaps surprisingly, did not diminish drug overdoses. Instead, it precipitated new risks and harms. We have since learned that reducing or discontinuing prescription opioids increases the risk of overdose, overdose deaths, and suicides, especially when tapering is forced or rapid [22-25]. Thus, more thought and research are needed to reduce these harms and improve patient outcomes.

It can be uniquely challenging to taper the opioids of patients with chronic pain. Patients take opioids to manage their pain and may develop some dependence over time. As a result, when patients reduce or stop their opioids, they may experience *both* increases in pain and craving, both of which are negative affective states [8, 26]. A recent cohort study highlighted The difficulty of tapering, which showed 21,515 tapering events in 19,377 patients reported adverse incidence rates. Ten other *retrospective* studies documented similar tapering-related risks, but no prospective studies exist. Despite its difficulty and risks, there are important benefits to tapering. Notably, over two-thirds of patients who initiate tapering sustain long-term dose reductions (> 16 months) [27], demonstrating the potential usefulness of this approach. However, the risk-reward profile may dampen physicians’ and patients’ enthusiasm for undertaking opioid tapering [28-31], which has been discussed extensively in the clinical community.

Patients’ and physicians’ enthusiasm for tapering may be partially uncovered by better understanding the patients’ opinions and attitudes toward tapering. These opinions and attitudes can be used to drive research on patient-centered tapering paradigms and to facilitate evidence-based tapering. Here, we aimed to take a first step in this direction. In this survey study, we queried opioid tapering attitudes, expectations, and reservations of chronic pain patients who have been using opioids to manage their pain for extended time periods. We focused on patients with chronic back pain because it is the most common homogeneous pain condition for which long-term opioids are typically prescribed [32] and is a leading cause of work-related disability [33]. Our results indicate an ambivalence by the patients surveyed. Most patients are highly interested in participating in opioid tapering research, with the caveat that their pain is controlled.

## Methods

One thousand participants (50% Female) from Prolific.com, an online research platform, were pre-screened for chronic pain and opioid use from their database of potential research participants located in the United States. Of those, 161 participants met our inclusion criteria of having chronic pain for more than six months and having been on opioid medication for more than six months. One hundred fifty pre-screens were invited to participate in the main study, which consisted of questions about their pain, opioid medications, desire to participate in a study to taper their opioid dose, as well as what tapering side effects are most important for them to control. Of the 150 invited, 127 consented to participate and completed the questionnaire.

Ordinal regression was performed to understand the dependence of the participant’s level of interest in tapering and the importance of controlling pain intensity when tapering their dose, on pain intensity, on the amount of opioid consumed (morphine milligram equivalent per day, MME), pain duration, and opioid use duration.

## Results

One hundred twenty-seven chronic pain patients (53.5% female) participated in our survey. They have an average pain duration of 13.5 years and have been on opioids for an average of 10.3 years. 76% of the participants are on Hydrocodone or Oxycodone (35.7%, 34.9%) with a mean MME of 56.2 mg (Table 1), and the majority is “very” to “extremely” interested in opioid tapering (30%, 36%) (Figure 1). 76% of the participants indicated that controlling pain intensity is “very” or “extremely” important to them while tapering down their opioid dose (28%, 58%), followed by maintaining the ability to perform daily routine, with 33% indicating it is “very” important and 41% indicating it is “extremely” important, followed by controlling mood, then opioid cravings (Figure 1).

**Table 1:** Participant’s demographics, pain, and opioid data.

| Variable | All Patients (n = 127) |
| --- | --- |
| **Sex** |  |
| Female | 68 (53.5%) |
| Male | 59 (46.5%) |
| **Mean Age (Years)** | 44.2 ± 13.0 |
| **Education** |  |
| Current Student | 16 (12.6%) |
| Not Currently a Student | 79 (62.2%) |
| **Employment** |  |
| Full-time employee | 59 (46.5%) |
| Not in paid work | 13 (10.2%) |
| Part-time employee | 10 (7.9%) |
| Unemployed and job seeking | 5 (3.9%) |
| **Race** |  |
| White | 109 (85.8%) |
| Black/African-American | 6 (4.7%) |
| Asian | 1 (0.8%) |
| American Indian/Alaska Native | 2 (1.6%) |
| More than one race | 8 (6.3%) |
| Declined to answer | 1 (0.8%) |
| **Mean Pain Duration (Years)** | 13.5 ± 9.6 |
| **Mean Opioid Use Duration (Years)** | 10.3 ± 8.2 |
| **Mean MME (mg)** | 56.2 ± 106.3 |
| **Medication** |  |
| Hydrocodone | 45 (35.7%) |
| Oxycodone | 44 (34.9%) |
| Tramadol | 16 (12.7%) |
| Morphine | 6 (4.8%) |
| Buprenorphine | 5 (4.0%) |
| Methadone | 4 (3.2%) |
| Codeine | 3 (2.4%) |
| Diacetylmorphine | 1 (0.8%) |
| Fentanyl patch | 1 (0.8%) |
| Tapentadol | 1 (0.8%) |

**Figure 1:**
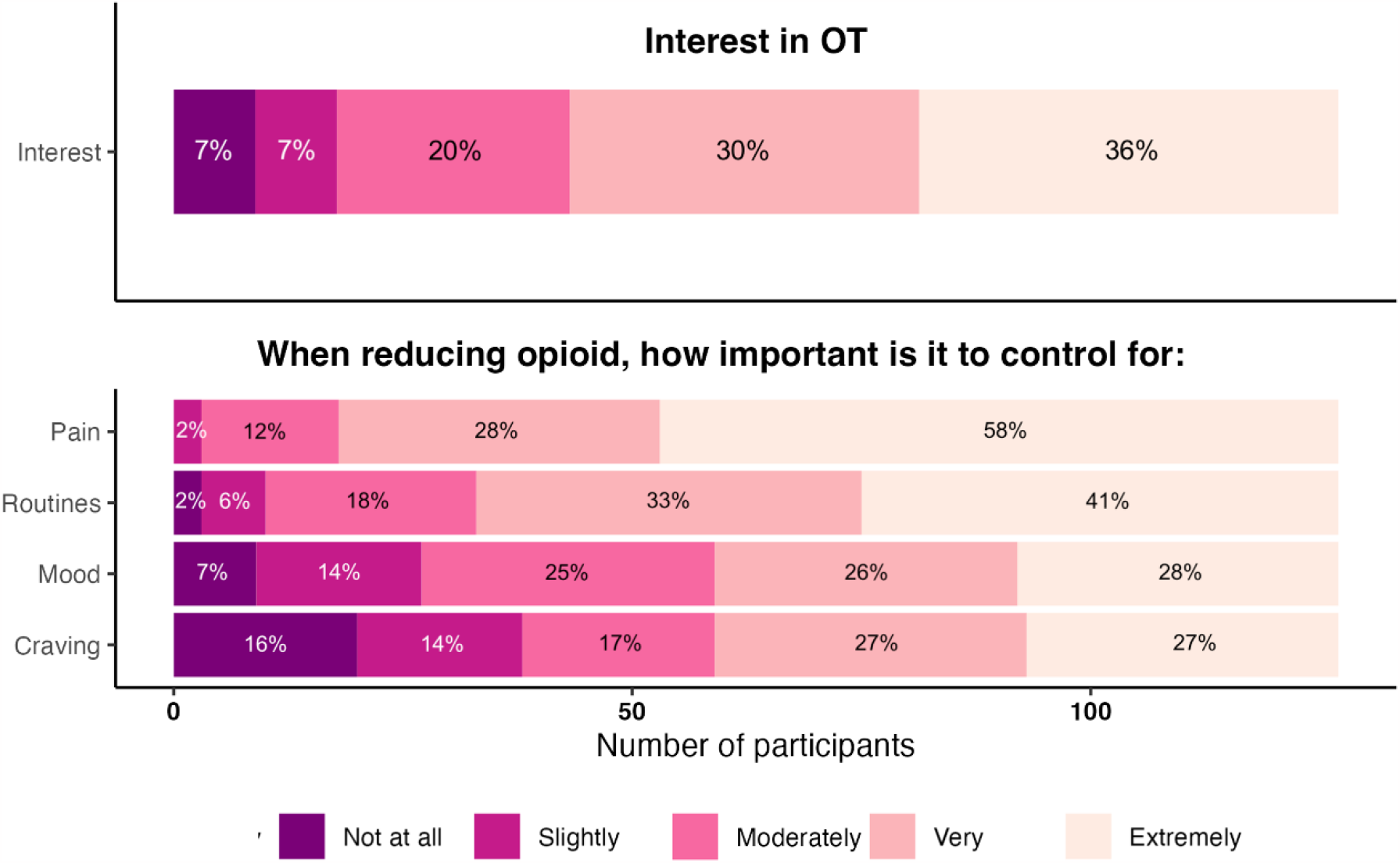
Interest and worries in opioid tapering. (A) 66% of the 127 participants are “very” or “extremely” interested in opioid tapering, with the majority being “extremely” interested. (B) Controlling pain intensity is the major worry for participants, with 76% indicating that it is “very” or “extremely” important. The second important worry is maintaining the ability to perform daily routines, with 74% indicating it is “very” or “extremely” important.

Ordinal regressions (Figure 2) show that the likelihood of being extremely interested in opioid tapering increases with increased pain intensity (*p=0*.*082*), becoming the most likely choice for participants with pain intensity greater than or equal to 6/10. Participants with pain intensity greater than 4/10 are more likely to choose “very” or “extremely” interested in opioid tapering (Figure 2A). However, we see no evidence of this likelihood being affected by pain duration (*p=0*.*98*) (Figure 2B). We also observe that regardless of opioid dose, the likelihood of being extremely interested in opioid tapering is highest; however, there is neither evidence that interest is dependent on dose (*p=0*.*59*) (Figure 2C), nor on opioid use duration (*p=0*.*63*) (Figure 2D).

**Figure 2:**
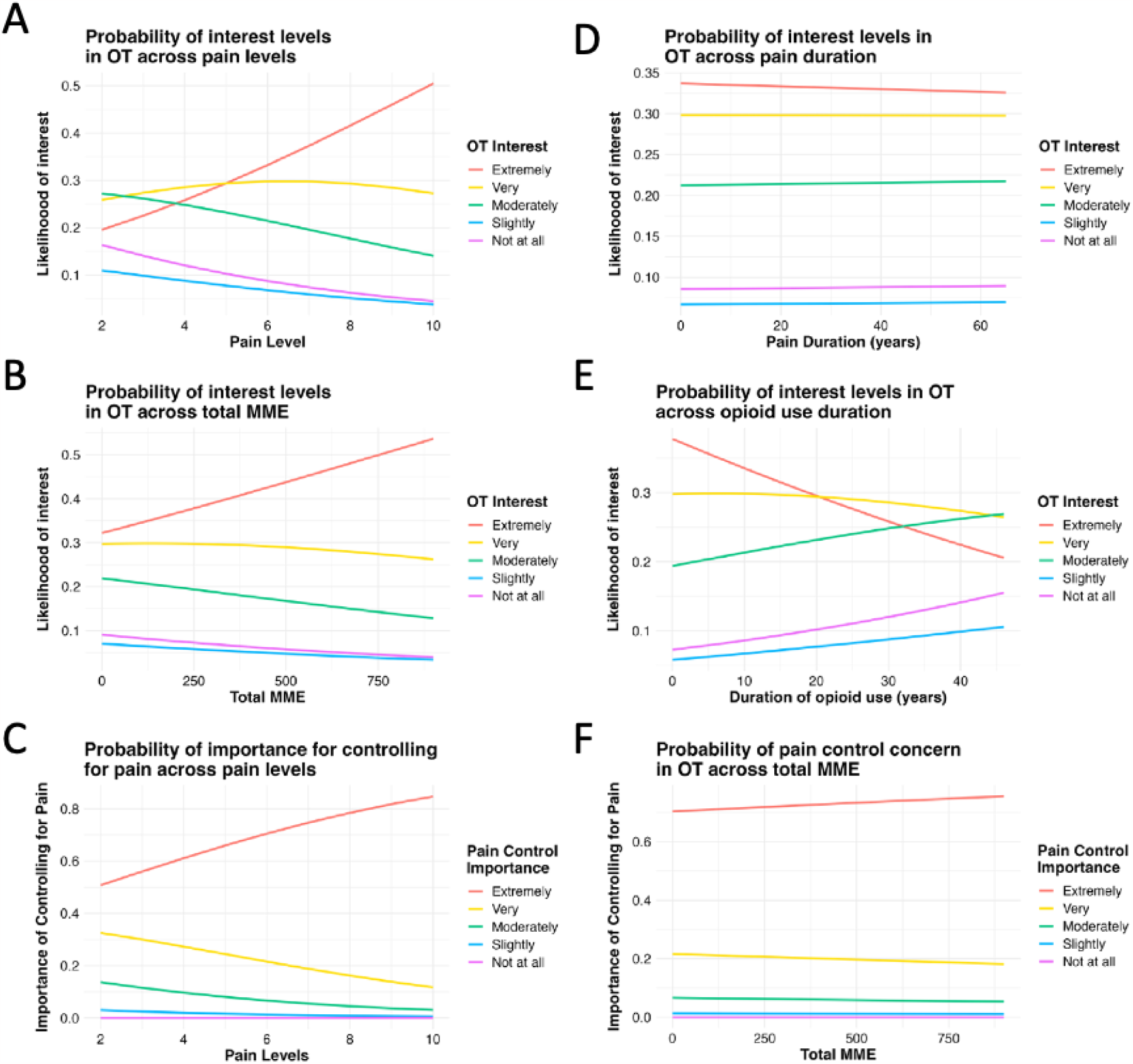
Relationship between patient pain level and medication MME, and interest in opioid tapering and importance of controlling pain. (A) The likelihood of being extremely interested in opioid tapering increases with increased pain intensity, becoming the most likely choice for participants with pain intensity greater than or equal to 6/10. Participants with pain intensity greater than 4/10 are more likely to choose “very” or “extremely” interested in opioid tapering. (B). (C) Regardless of their opioid dose, the likelihood of being highly interested in opioid tapering is highest, and this likelihood increases for higher MME doses. (D) The likelihood of “controlling pain” being extremely important for opioid tapering is always highest and is not dependent on MME.

With regards to the importance of controlling pain when tapering, we observe an increase in the probability of being “extremely” interested in controlling pain with increased pain levels (*p=0*.*0675*) (Figure 2E), but no evidence of it being affected by the opioid dose (*p=0*.*856*) (Figure 2F).

We also note that participants to whom controlling pain intensity is “very” or “extremely” important are no less interested in opioid tapering than those who reported that controlling pain is “moderately” or “slightly” important. 61% of the participants for whom pain control is extremely important and 83% of those for whom it is very “important” are “very” to extremely” interested in pain, whereas, for those where pain control is “moderately” or “slightly” important the percentages are 60 and 34 (Figure 3).

**Figure 3:**
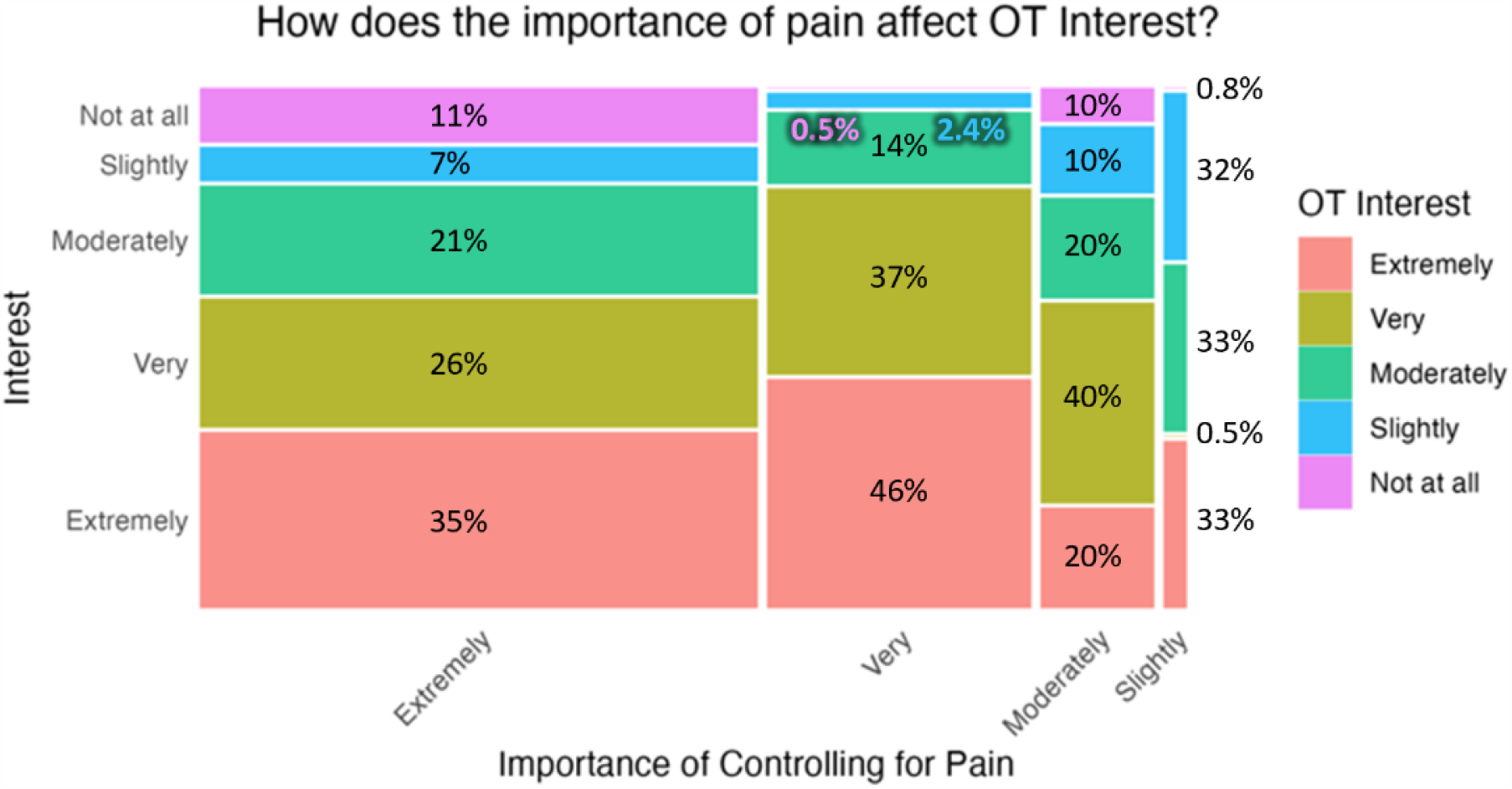
Interest in opioid tapering and the importance of controlling pain. Participants who are “very” or “extremely” worried about controlling pain intensity are not less interested in opioid tapering than those who are “moderately” or “slightly” worried about controlling pain intensity (61% and 83% vs 60% and 34%).

## Discussion

Our results indicate that the chronic pain patients on long-term opioid therapy, surveyed from the Prolific website, were highly interested in participating in opioid tapering research. However, they emphasized the importance of controlling their pain during tapering. We also observe that their interest does not seem to be dependent on pain duration, opioid dose, or duration of opioid consumption. With regards to the level of importance of controlling pain while tapering, our results show that it increases with increased pain intensity but does not seem to be affected by opioid dose. We also observe that the participants who are most worried about controlling pain intensity are not less interested in opioid tapering than those who have placed a lower priority on controlling pain intensity. Although the participants emphasize control of pain during tapering, over 50% acknowledge the need to control their mood and opioid craving. Thus, the latter group is cognizant of their opioid use dependence, and they are probably the group most vulnerable to harm following opioid tapering. This survey serves as an important first step in the systematic study of opioid tapering in patients with chronic pain in a patient-centered fashion.

Surprisingly, this survey seems to be one of very few of its kind. Still, the observed results show consistencies within the minimal literature related to the topic. A very recent study used semi-structured interviews and analyzed patient and provider preferences about long-term opioid use continuation and discontinuation and non-opioid pain treatments at two US Veterans Affairs institutions [34]. The study included 28 patients and 15 providers. In contrast to providers, patients were uncertain of the utility of opioid tapering and stated that its harms would outweigh its benefits. Their main worry regarding tapering—like our results—was worsening their ongoing pain. Yet, we should emphasize that the two surveys come from very different communities. Another study of group and individual interviews of 21 adults with back pain undergoing tapering [35] revealed the complexity of attitudes and decisions, as well as the dynamic nature of the needs of patients undergoing tapering. Individual patients’ values influenced the extent of their interest in tapering. Importantly and consistent with our results, fear primarily affected patients’ willingness to taper. Particularly fears involving the possibility of worsening pain and withdrawal due to decreased opioid dose, suggesting a relationship between willingness to taper and opioid duration. Our results showed no evidence of such a relationship. Many of the participants indicate that it is common before social activities for patients to take their opioids to reduce their pain for a predictable duration. Losing control over when to lower their pain, which allows them the ability to stay socially engaged and to have a daily routine, may also contribute to their fear of tapering. This is a hypothesis that our survey supports, with 74% of the respondents indicating that being able to perform daily routines is “very” or “extremely” important.

It is important to note that a recent study that closely matches our approach and the number of participants queried [36] concluded that “patients with lower pain severity, shorter duration of pain, and higher concerns about opioids may be a prime target from a motivation standpoint for interventions addressing opioid tapering and discontinuation.” Our results show that interest in tapering increases, rather than decreases, with pain intensity. This difference may be due to differences in sample characteristics, particularly the employment status, where almost half of our sample were full-time employed, whereas, in the sample reported in [36], only 18.5 were full-time employed. There may be other important differences in the populations surveyed by us and by [36], which remain unknown.

## Conclusions

Our results show that most surveyed chronic pain patients on long-term opioid treatment are highly interested in opioid tapering, with controlling pain being their primary concern, followed closely by the ability to perform daily routines. Some of these results agree with what few have been published, others do not, possibly due to the different biases of the samples in each study. However, across these studies, concern about pain control seems to be a dominant theme. The latter is hard to understand, especially as multiple studies indicate the minimal efficacy of long-term opioid use in controlling chronic pain. A recent study in a large group of subjects indicates that even post-tapering, there is no change in pain or pain interference [37]. Taken together, the results suggest that such patients become dependent on a cycle of transient pain relief and mood normalization with each exposure to their opioids, enabling daily functioning. It is likely that such cyclic changes in pain and mood are a consequence of the neuropharmacology of long-term opioid exposure, which, once established, becomes very hard to abrogate. To develop successful and humane tapering protocols, it is imperative that a larger and more coordinated effort be invested in studying the therapeutic preferences, needs, and fears of patients on long-term opioid treatment who are considering tapering their opioid consumption and address these issues individually and dynamically during the process of tapering.

## Data Availability

All data produced in the present study are available upon reasonable request to the authors.

